# NT-proBNP Thresholds for Early Heart Failure Detection in Asian Patients With Type 2 Diabetes

**DOI:** 10.64898/2026.02.27.26347295

**Authors:** Tai-Shuan Lai, Chia-Ling Tseng, Cho-Kai Wu, Liang-Ting Chiang, Yong-Chen Chen, Wan-Lun Hsu

## Abstract

**Background:** Heart failure (HF) is an increasingly common complication among patients with type 2 diabetes (T2D), yet its early detection remains challenging, especially in those with concomitant chronic kidney disease (CKD). NT-proBNP is a key biomarker for diagnosing and prognosticating HF, but its reference thresholds are influenced by renal function, age, and ethnicity. Current guideline cutoffs, largely derived from Western populations, may not apply to Asian patients.

**Methods:** This retrospective cohort study included 10,587 adults with T2D who underwent NT-proBNP testing between 2006 and 2021 at the National Taiwan University Hospital. Patients with prior HF were excluded. Generalized additive models identified NT-proBNP thresholds associated with HF hospitalization, and Kaplan–Meier analysis validated outcome separation. Subgroup analyses were stratified by age, sex, body mass index (BMI), and estimated glomerular filtration rate (eGFR).

**Results:** During a mean follow-up of 3.5 years, 1,892 (17.9%) patients were hospitalized for HF. NT-proBNP levels of 179 pg/mL (outpatient) and 728 pg/mL (emergency) marked inflection points for rising event risk (log-rank *p* < 0.0001). Age-specific analyses showed progressive increases in optimal thresholds: from 85 (<50 years old), 150 (50–74 years old) and 290 pg/mL (≥75 years old) in outpatients, and from 310, 600 and 1,165 pg/mL, respectively, in emergency settings. In the BMI-stratified analysis, NT-proBNP thresholds demonstrated an inverse relation with BMI. Considering renal function, the optimal cutoffs were 100, 310, and 935 pg/mL for eGFR > 60, 30–60, and < 30 mL/min/1.73 m², respectively; in the emergency cohort, the corresponding thresholds were 290, 835, and 3,905 pg/mL.

**Conclusions:** This large Asian cohort defines setting- and renal function-specific NT-proBNP thresholds for predicting HF hospitalization in patients with T2D. The lower optimal cutoffs compared with Western guidelines highlight the need for ethnicity-adjusted diagnostic criteria to improve early identification and risk stratification of HF in clinical practice.

**What is new?:**

- In a large real-world Asian cohort of patients with type 2 diabetes, we identified setting-specific NT-proBNP thresholds (179 pg/mL outpatient; 728 pg/mL emergency) associated with heart failure hospitalization risk.
- Age-, BMI-, and kidney function–stratified cutoffs revealed substantial heterogeneity in optimal NT-proBNP thresholds.
- Compared with guideline-recommended values, Asian-specific thresholds were consistently lower (∼30–40%), supporting ethnic differences in natriuretic peptide biology.
- A generalized additive model (GAM) captured nonlinear biomarker–risk relationships, enabling data-driven and clinically interpretable cutoff identification.

**What are the clinical implications?:**

- Use of ethnicity- and context-specific NT-proBNP thresholds may improve early detection of heart failure in Asian patients with type 2 diabetes.
- Incorporating kidney function and BMI into NT-proBNP interpretation enhances risk stratification, particularly in patients with CKD.
- Reliance on Western guideline cutoffs may underestimate heart failure risk in Asian populations.
- These findings support a precision medicine approach to biomarker interpretation and highlight the need for population-specific guideline refinement.

## Introduction

Type 2 diabetes (T2D) has become an increasingly common non-communicable disease, estimated to affect 380 million people worldwide.^1^ Patients with T2D are at an increased risk of cardiovascular diseases and associated clinical complications.^2^ The incidence of myocardial infarction and stroke has declined during the past few decades, whereas that of heart failure has gradually increased. Heart failure affects up to 22% of people with diabetes.^3^ Compared with patients without diabetes, patients with diabetes have an increased risk of heart failure of up to 85%, with a risk ratio of 1.85 (95% CI: 1.51–2.28).^4^ Among cardiovascular complications, heart failure is one of the earliest to occur.^5^

However, predicting the risk of heart failure in people with T2D can be challenging. Many clinical risk factors are associated with increased risk of heart failure in people with T2D, such as hypertension, obesity, atrial fibrillation, and aging.^6,7^ Furthermore, non-specific symptoms of heart failure, such as breathlessness, ankle swelling, and fatigue, are often confused with chronic lung disease, liver cirrhosis, or chronic kidney disease (CKD), complicating the timely diagnosis of heart failure.^8^ Biomarkers such as NT-proBNP help in the early diagnosis of heart failure, especially in patients with diabetes.

NT-proBNP is a cardiac-specific biomarker, which is cleaved by furin from proBNP. When the myocardial wall is stressed, NT-proBNP will be stimulated and secreted.^9^ Increased NT-proBNP concentration serves as an indication of heart failure and correlates with symptom severity. Compared with BNP, another cleavage from proBNP, NT-proBNP is biologically inactive: it is stable in room temperature and has no physiological functions.^10^ Therefore, NT-proBNP serves as a reliable and objective tool for screening patients at risk of heart failure, diagnosing acute and chronic heart failure, and disease monitoring, as a prognostic factor of heart failure.^11–13^

The Heart Failure guideline from the European Society of Cardiology (ESC) 2021 recommends an NT-proBNP level < 125 pg/mL as the threshold for ruling out chronic heart failure. This 125 pg/mL cutoff in primary care offers high sensitivity (94.6%) but moderate specificity (50%). For acute heart failure in emergency or hospital settings, a threshold of NT-proBNP < 300 pg/mL effectively excludes acute heart failure.^12^ Notably, several clinical contexts can influence NT-proBNP values or their interpretation, including age, renal function, and obesity. For instance, in chronic heart failure, the threshold of NT-proBNP is suggested at > 250 pg/mL and > 500 pg/ml for patients aged 50–74 years and over 75 years, respectively.^7^ Meanwhile, studies have not provided supportive evidence for cutoff values for patients at different stages of renal function and with different levels of BMI. Furthermore, these guidelines are based on studies with White Europeans, and evidence for Asian populations is lacking. Thus, we aimed to conduct a hospital-based study to set up the threshold of NT-proBNP for acute and chronic heart failure in general and specified conditions.

## Methods

### Data Sources

We conducted this hospital-based retrospective cohort study using claims data from the Integrated Medical Database of the National Taiwan University Hospital (NTUH-IMD).^14^ The database includes structured, semi-structured, and unstructured data, such as demographic information, diagnoses, medical orders, laboratory tests, visit records, prescriptions, and deaths. Death data are further linked from the National Death Registry. This study was approved by the Research Ethics Committee of NTUH and complied with the Declaration of Helsinki (approval number: 202402081RIFE). The data underlying this study are not publicly available owing to patients’ privacy but are available from the corresponding author upon reasonable request and with approval from the Institutional Review Board.

### Study Population

In this cohort study, we included patients with T2D aged >18 years who underwent NT-proBNP testing in outpatient and emergency settings between January 1, 2006, and December 31, 2021. We defined T2D using codes from the International Classification of Disease, 9th and 10th revisions (ICD-9, ICD-10), as listed in Supplementary Table 1. The inclusion criteria were two outpatient diabetes diagnosis within one year, or one outpatient diagnosis of diabetes and outpatient prescription for hypoglycemic drugs and inpatient diagnosis of diabetes or prescription of hypoglycemic drugs during hospitalization. We set the index date of enrollment in the cohort as the date of the first NT-proBNP testing. We excluded patients with type 1 diabetes, a history of heart failure hospitalization prior to the index date, logically inconsistent dates (e.g., NT-proBNP testing date after December 31, 2021), and a follow-up period of less than one month. Our final sample for analyses included 10,587 patients. The study flowchart is presented in Figure 1.

**Figure 1.**
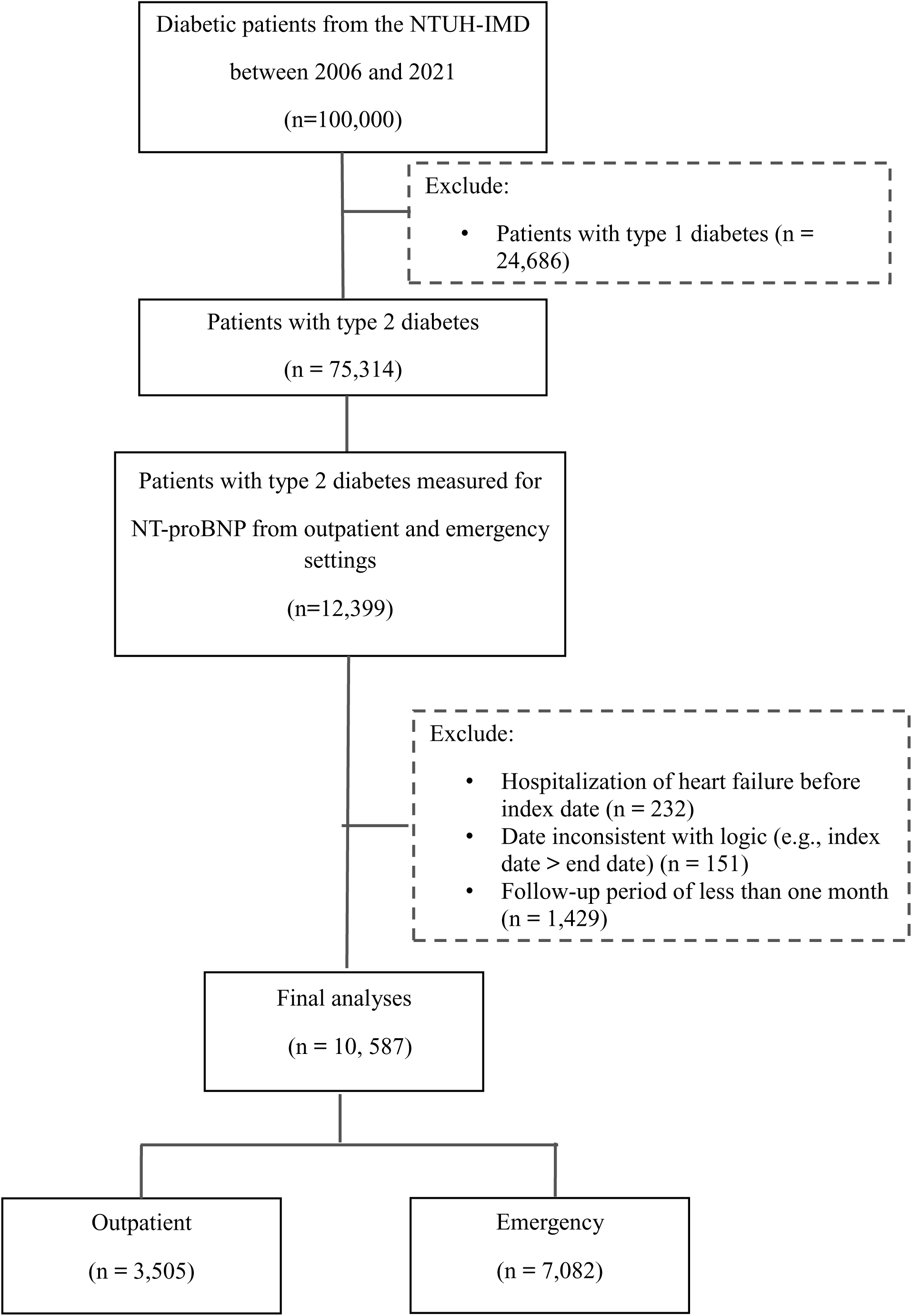
Patient selection flow chart.

### Study Definitions

We defined baseline comorbidities, such as hypertension (HTN), cerebrovascular disease (CVD), and coronary heart disease (CHD), using ICD codes (Supplementary Table 1). We measured Serum NT-proBNP level by electrochemiluminescence immunoassay. The reference range of NT-proBNP was < 125 pg/mL. We used colorimetry to measure serum creatinine level and turbidimetric immunoassay to measure urine microalbumin. The reference range of microalbumin was < 30 mg/L. We calculated the estimated glomerular filtration rate (eGFR) using the Chronic Kidney Disease Epidemiology Collaboration (CKD-EPI) equation.^15^ We assessed urine protein levels using dipstick urinalysis and categorized them as negative or positive based on semi-quantitative results. Negative results included “negative” and “-“ (no detectable protein); positive results included any levels from 1+ to 4+.

### Study Outcome

The observation endpoint of the study was heart failure hospitalization, defined using ICD-9-CM code 428.xx and ICD-10-CM code I50.xx. Patients were followed until the occurrence of heart failure hospitalization, death, or the study end date (December 31, 2021), whichever came first.

### Statistical Analyses

The continuous variables are shown as mean value ± standard deviation (SD) (or median with interquartile range, IQR), and categorical variables are presented as counts and percentages. We performed log transformation for skewed data. We used Kaplan–Meier curves and log-rank tests to visualize the data.

### NT-proBNP Cutoff Point

To determine the NT-proBNP cutoff for heart failure hospitalization, we adopted a generalized additive model (GAM) with splines for analyzing the relation between NT-proBNP levels and heart failure risk. This approach permits adjustments for possible nonlinear effects of continuous variables. To show the effect of NT-proBNP on risk of heart failure, we plotted a function curve with values of the logs of odds ratios. We centered the curve to set an average of zero over the range of the data, with approximate pointwise 95% confidence intervals (CIs) shaded; we also calculated for sensitivity and specificity.

All analyses were performed using SAS 9.4 (SAS Institute Inc., North Carolina, USA) and R software, version 4.4.1 (Free Software Foundation, Inc., Boston, MA, U.S.A.). A two-sided p value < 0.05 was considered statistically significant.

## Results

From nearly 80,000 adults with T2D, we identified a cohort of 10,587 individuals who underwent NT-proBNP testing, including 3,505 from outpatient and 7,082 from emergency settings (Figure 1). Table 1 summarizes their demographic and clinical characteristics.

**Table 1.**
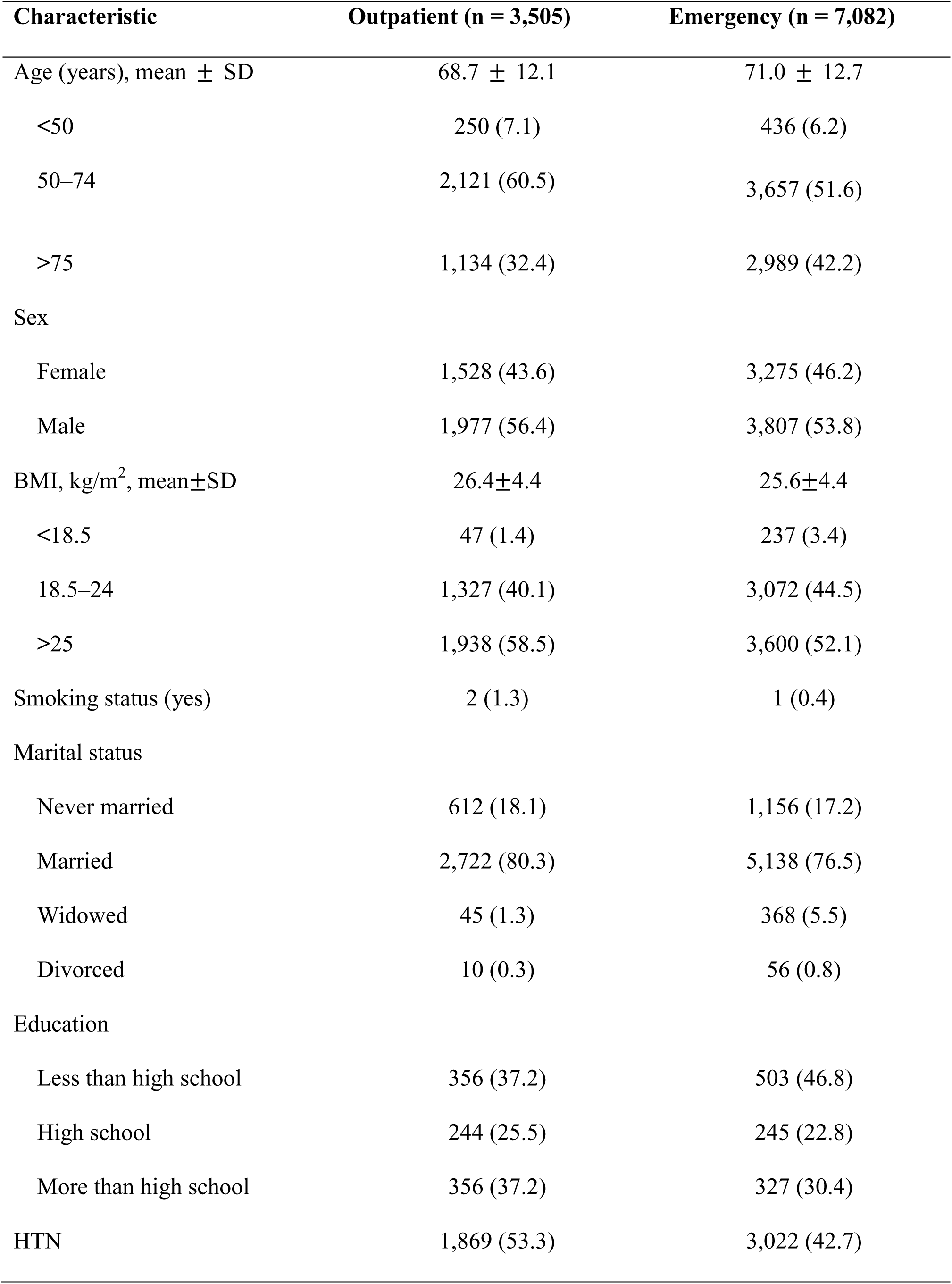
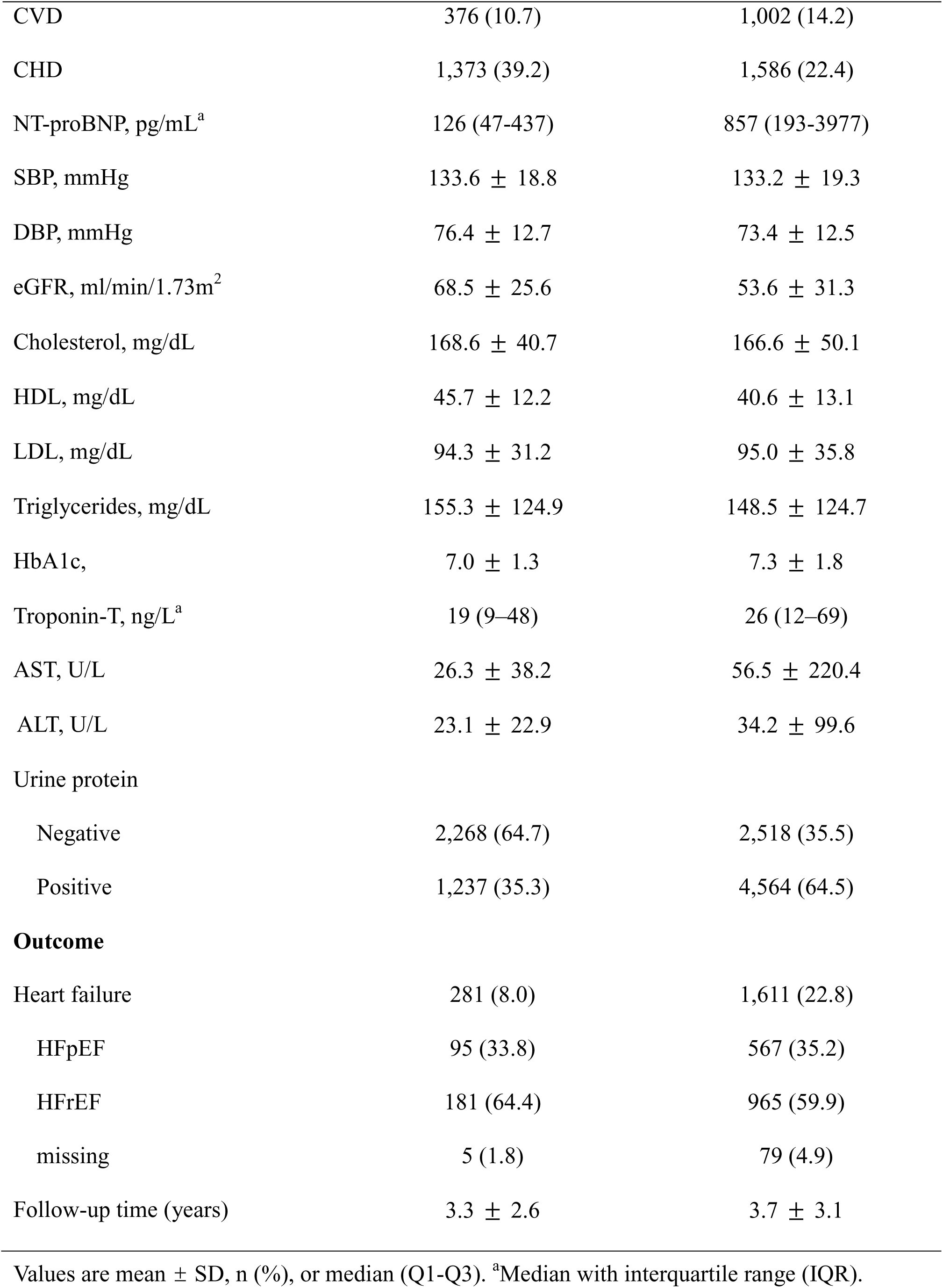
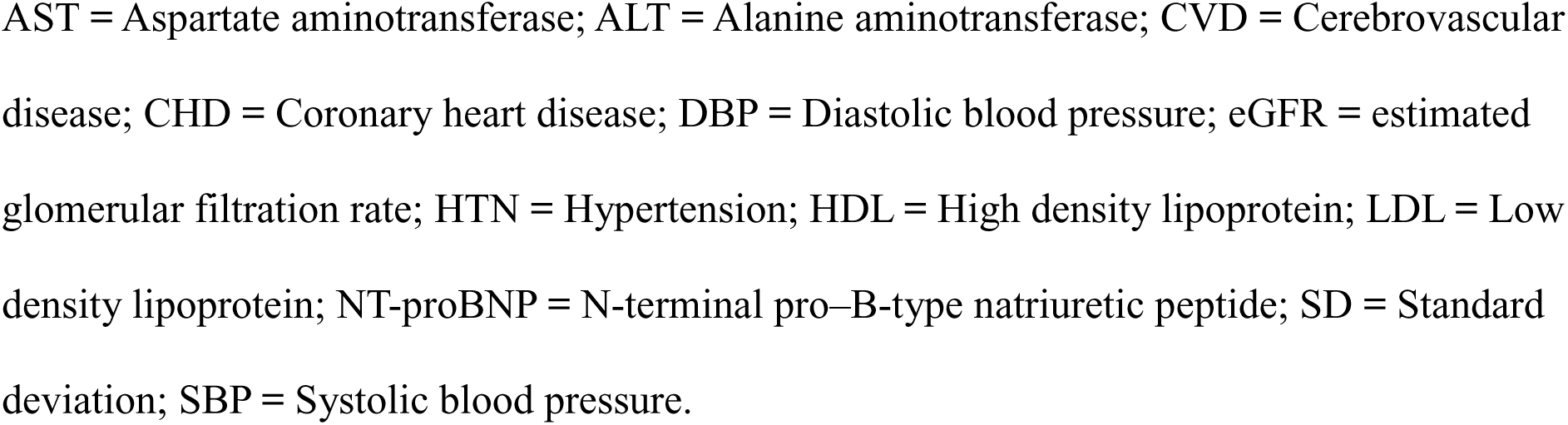
Patient characteristics of the outpatient and emergency cohorts.

| Characteristic | Outpatient (n = 3,505) | Emergency (n = 7,082) |
| --- | --- | --- |
| Age (years), mean ± SD | 68.7 ± 12.1 | 71.0 ± 12.7 |
| <50 | 250 (7.1) | 436 (6.2) |
| 50–74 | 2,121 (60.5) | 3,657 (51.6) |
| >75 | 1,134 (32.4) | 2,989 (42.2) |
| Sex |  |  |
| Female | 1,528 (43.6) | 3,275 (46.2) |
| Male | 1,977 (56.4) | 3,807 (53.8) |
| BMI, kg/m<sup>2</sup>, mean±SD | 26.4±4.4 | 25.6±4.4 |
| <18.5 | 47 (1.4) | 237 (3.4) |
| 18.5–24 | 1,327 (40.1) | 3,072 (44.5) |
| >25 | 1,938 (58.5) | 3,600 (52.1) |
| Smoking status (yes) | 2 (1.3) | 1 (0.4) |
| Marital status |  |  |
| Never married | 612 (18.1) | 1,156 (17.2) |
| Married | 2,722 (80.3) | 5,138 (76.5) |
| Widowed | 45 (1.3) | 368 (5.5) |
| Divorced | 10 (0.3) | 56 (0.8) |
| Education |  |  |
| Less than high school | 356 (37.2) | 503 (46.8) |
| High school | 244 (25.5) | 245 (22.8) |
| More than high school | 356 (37.2) | 327 (30.4) |
| HTN | 1,869 (53.3) | 3,022 (42.7) |
| CVD | 376 (10.7) | 1,002 (14.2) |
| CHD | 1,373 (39.2) | 1,586 (22.4) |
| NT-proBNP, pg/mL<sup>a</sup> | 126 (47-437) | 857 (193-3977) |
| SBP, mmHg | 133.6 ± 18.8 | 133.2 ± 19.3 |
| DBP, mmHg | 76.4 ± 12.7 | 73.4 ± 12.5 |
| eGFR, ml/min/1.73m<sup>2</sup> | 68.5 ± 25.6 | 53.6 ± 31.3 |
| Cholesterol, mg/dL | 168.6 ± 40.7 | 166.6 ± 50.1 |
| HDL, mg/dL | 45.7 ± 12.2 | 40.6 ± 13.1 |
| LDL, mg/dL | 94.3 ± 31.2 | 95.0 ± 35.8 |
| Triglycerides, mg/dL | 155.3 ± 124.9 | 148.5 ± 124.7 |
| HbA1c, | 7.0 ± 1.3 | 7.3 ± 1.8 |
| Troponin-T, ng/L<sup>a</sup> | 19 (9–48) | 26 (12–69) |
| AST, U/L | 26.3 ± 38.2 | 56.5 ± 220.4 |
| ALT, U/L | 23.1 ± 22.9 | 34.2 ± 99.6 |
| Urine protein |  |  |
| Negative | 2,268 (64.7) | 2,518 (35.5) |
| Positive | 1,237 (35.3) | 4,564 (64.5) |
| **Outcome** |  |  |
| Heart failure | 281 (8.0) | 1,611 (22.8) |
| HFpEF | 95 (33.8) | 567 (35.2) |
| HFrEF | 181 (64.4) | 965 (59.9) |
| missing | 5 (1.8) | 79 (4.9) |
| Follow-up time (years) | 3.3 ± 2.6 | 3.7 ± 3.1 |

Values are mean ± SD, n (%), or median (Q1-Q3). <sup>a</sup>Median with interquartile range (IQR).
AST = Aspartate aminotransferase; ALT = Alanine aminotransferase; CVD = Cerebrovascular disease; CHD = Coronary heart disease; DBP = Diastolic blood pressure; eGFR = estimated glomerular filtration rate; HTN = Hypertension; HDL = High density lipoprotein; LDL = Low density lipoprotein; NT-proBNP = N-terminal pro-B-type natriuretic peptide; SD = Standard deviation; SBP = Systolic blood pressure.

### NT-proBNP Cutoff Point by GAM

We used a GAM to identify the cutoff of NT-proBNP levels associated with the risk of heart failure hospitalization (Figure 2). We defined log-transformed NT-proBNP values of 5.19 (approximately 179.47 pg/mL on the exponential value) and 6.59 (approximately 727.78 pg/mL on the exponential value) as the outpatient and emergency cutoff values, respectively. The sensitivity for predicting heart failure hospitalization in the outpatient and emergency settings was 78.29% and 79.39%, respectively. Kaplan–Meier survival plots showed that the different NT-proBNP level group was significantly associated with the risk of heart failure hospitalization, with the log-rank test *p* < 0.0001 (Figure 3).

**Figure 2.**
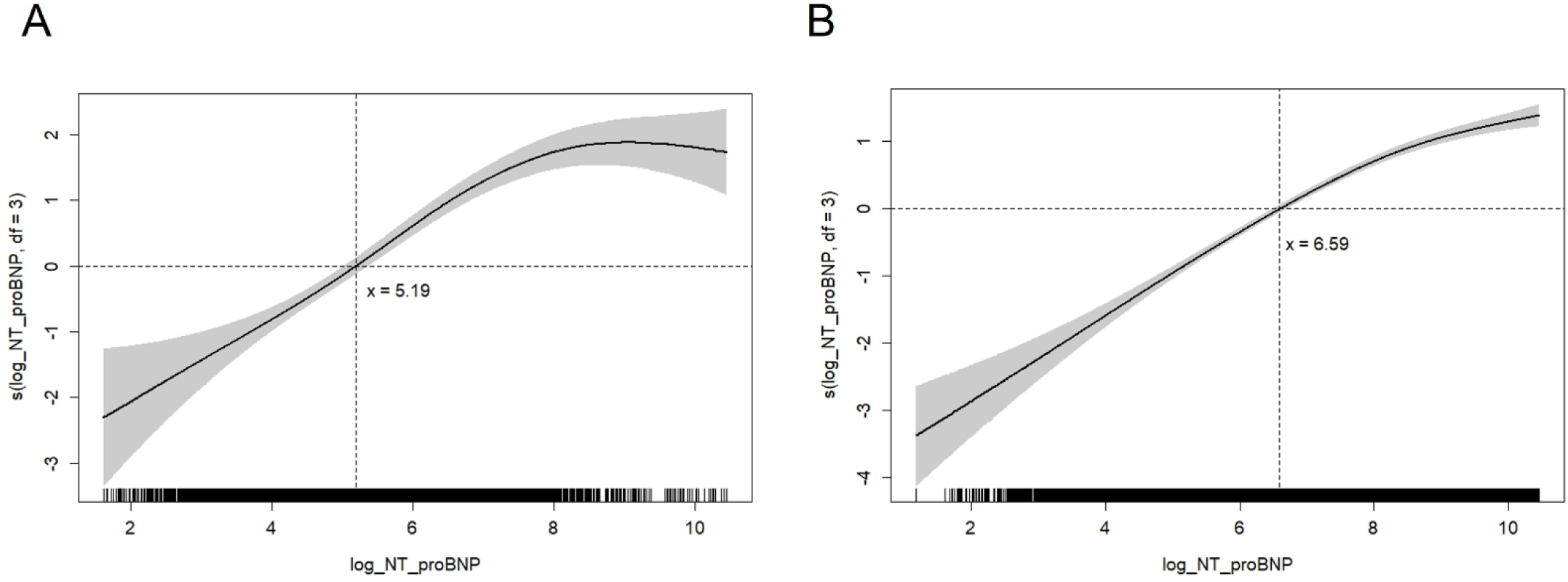
Function curve representing the smooth terms of the generalized additive model with splines for analyzing the relation between log-transformed NT-pro BNP and risk of heart failure hospitalization. The plot illustrates the nonlinear association between log-transformed NT-proBNP and log of odds ratios for heart failure hospitalization, with 95% confidence intervals shaded in gray. (A) In the outpatient cohort, the cutoff was set at a log-transformed NT-proBNP value of 5.19 (approximately 179.47 pg/mL). (B) In the emergency cohort, the cutoff was set at a log-transformed NT-proBNP value of 6.59 (approximately 727.78 pg/mL). NT-proBNP = N-terminal pro-B-type natriuretic peptide.

**Figure 3.**
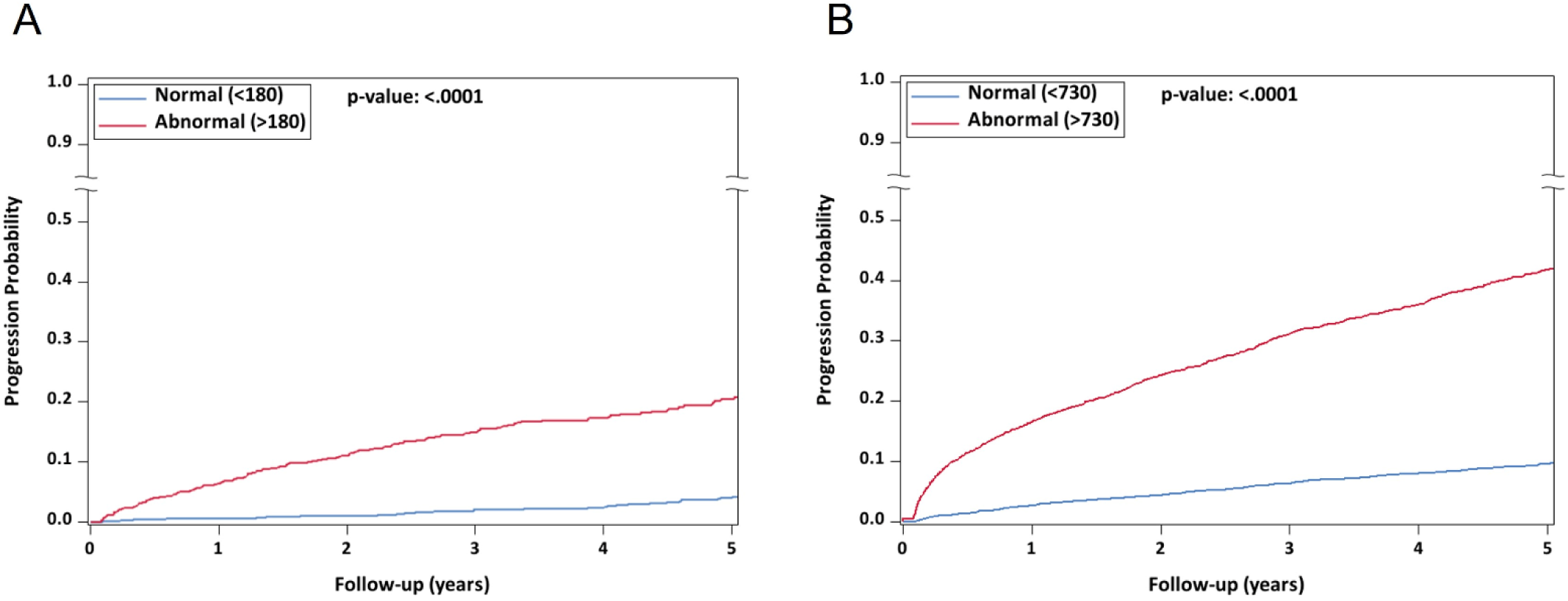
Cumulative incidence of heart failure hospitalization between different NT-proBNP level groups. Patients were divided into groups based on NT-proBNP cutoff. (A) In the outpatient cohort, patients with abnormal NT-proBNP had a significantly higher risk of hospitalization compared with those with normal NT-proBNP. (B) In the emergency cohort, patients with abnormal NT-proBNP showed a significantly higher risk than those with normal NT-proBNP. NT-proBNP = N-terminal pro-B-type natriuretic peptide.

### Cumulative Incidence of Heart Failure Hospitalization Stratified by Different Variables

Figure S1–S8 display the cumulative incidence of heart failure hospitalization, grouped by different variables in the outpatient and emergency cohorts. We generated these plots according to the exponential values from Table 2 as the grouping criterion. In the outpatient group, patients aged >75 years had a higher cumulative incidence of heart failure hospitalization compared with the other two age groups. Patients with BMI <18.5 and eGFR <30 also showed a higher risk, indicating that underweight and severely impaired renal function are key factors in the risk of heart failure hospitalization. We observed a similar pattern in the emergency cohort. Patients aged >75 years, with a BMI <18.5, and eGFR <30 consistently exhibited a higher cumulative incidence of heart failure hospitalization. The differences in cumulative incidence were more pronounced in the emergency cohort compared with the outpatient cohort.

**Table 2.** Identified cutoff points for different variables based on log-transformed and exponential values using the generative additive.

|  | Age (years) |  |  | Sex |  | BMI |  |  | eGFR |  |  |
| --- | --- | --- | --- | --- | --- | --- | --- | --- | --- | --- | --- |
|  | <50 | 50–74 | ≥75 | Female | Male | <18.5 | 18.5–24 | ≥25 | >60 | 30–60 | <30 |
| Outpatient | 85 | 150 | 290 | 175 | 180 | 1,000 | 205 | 170 | 100 | 310 | 935 |
| Emergency | 310 | 600 | 1,165 | 810 | 660 | 1,435 | 890 | 560 | 290 | 835 | 3,905 |

This table summarizes the cutoff values of NT-proBNP identified using a generalized additive model across different variables. Cutoff points were calculated based on both log-transformed and exponential values, and are presented according to age, sex, BMI, and eGFR for both outpatient and emergency cohorts. eGFR = estimated glomerular filtration rate.

## Discussion

In this large hospital-based retrospective cohort study utilizing real-world data from a tertiary medical center in Taiwan, we identified care setting-specific NT-proBNP thresholds that distinguished differences in cumulative incidence of heart failure hospitalization among patients with T2D. Using a GAM, we determined that NT-proBNP levels of 179 pg/mL for outpatients and 728 pg/mL for emergency cases corresponded to observable inflection points in the risk curve. These thresholds were further supported by Kaplan–Meier survival analysis, which revealed significant separation in heart failure hospitalization rates between groups stratified by these values (log-rank *p* < 0.0001). Notably, we also explored whether optimal NT-proBNP thresholds varied across subgroups defined by age, sex, BMI, and eGFR. The identification of distinct cutoff points across these clinical strata suggests that personalized interpretation of NT-proBNP may further enhance risk stratification.

To our knowledge, this is the largest study in a diabetic population to propose empirically derived, context- and subgroup-specific NT-proBNP thresholds using nonlinear modeling, offering a data-driven approach to support individualized cardiovascular risk assessment. In our study, the cutoff value of NT-proBNP in chronic heart failure is 179 pg/mL, which is slightly higher than the cutoff value of 125 pg/mL guided by the ESC; the cutoff value of NT-proBNP in acute heart failure is 728 pg/mL, which is much higher than the ESC cutoff value of 450 pg/mL.

Compared with the ESC guideline-recommended, age-stratified, rule-in thresholds for NT-proBNP (125, 250, and 500 pg/mL for <50, 50–74, and ≥75 years old, respectively), our data showed consistently lower cutoff values of 85, 150, and 290 pg/mL in the outpatient setting and 310, 600, and 1,165 pg/mL in the emergency setting. These thresholds maintain the same age-progressive pattern as the ESC recommendations but are approximately 30–40% lower across all age groups. We speculate that this difference may be partly attributable to the age distribution of our study cohort, which could have contributed to a higher overall cutoff, as relatively few participants in the cohort were under the age of 50. This difference likely reflects population-specific physiological and genetic factors influencing natriuretic peptide levels in East Asians, supporting the need for ethnicity-adjusted diagnostic criteria in heart failure evaluation. Ethnic variation in circulating NT-proBNP concentrations is a well-recognized phenomenon. Large multiethnic cohorts have consistently demonstrated that Asian populations exhibit substantially lower baseline NT-proBNP levels compared with White or European counterparts, even after adjustments for age, sex, BMI, and renal function.^16,17^ Population-based studies from Japan, Korea, and Singapore have further confirmed that reference ranges in East Asian populations are lower than those reported in Western cohorts, likely reflecting differences in ventricular wall stress, cardiac morphology, and genetic determinants of natriuretic peptide synthesis and clearance.^18,19^ Consequently, the lower NT-proBNP rule-in thresholds observed in our Taiwanese cohort are biologically plausible and consistent with regional data. These findings underscore the need for population-specific calibration of diagnostic cutoffs, rather than direct adoption of thresholds derived from European datasets, such as those used in the ESC guidelines.^20,21^

In patients with CKD or impaired renal function, NT-proBNP consistently reflects both cardiac and renal interactions across diverse populations. In the CRIC study,^22^ higher NT-proBNP levels in over 3,000 CKD participants without heart failure are strongly associated with left ventricular hypertrophy (LVH) and systolic dysfunction, improving risk classification when added to clinical models. Similarly, in outpatients with CKD in the US, deFilippi et al. showed that elevated NT-proBNP independently indicates underlying coronary artery disease and LVH, suggesting its rise represents subclinical cardiac injury rather than reduced clearance.^23^ In a European heart failure study, combining NT-proBNP > 4647 pg/mL with eGFR < 60 mL/min/1.73 m² could best predict the 60-day mortality across multicenter cohorts, defining a biomarker-based “cardiorenal syndrome.”^24^ Among Asian cohorts, Chen et al. demonstrated in 2,087 Chinese patients with coronary artery disease that NT-proBNP increases with declining renal function, with eGFR-specific cutoffs (179, 1,443, 3,478 pg/mL) improving prognostic accuracy.^25^ Another cohort study from China further showed that NT-proBNP correlates inversely with eGFR (r = -0.525), and the optimal NT-proBNP cutoff for diagnosing heart failure increases from 481 pg/mL in patients without CKD to 3,314 pg/mL in patients with CKD, demonstrating that diagnostic thresholds must be adjusted according to renal function stage.^26^ In Thai patients, Srisawasdi et al. reported that while both BNP and NT-proBNP rise with renal deterioration, NT-proBNP is more sensitive, and the NT-proBNP/BNP ratio remains the most stable indicator across renal stages and cardiac function.^27^ Our study strongly supports previous findings from both Western and Asian populations and further provides stage-specific NT-proBNP cutoff values for CKD. These values can help predict heart failure outcomes and guide clinical assessment in patients with varying CKD severity.

This study has several key strengths. First, in this long-term cohort with an average follow-up of more than three years, we demonstrated that NT-proBNP elevation precedes the clinical onset of heart failure in patients with T2D, supporting its role as an early predictor for subclinical cardiac dysfunction. Second, we applied a GAM to flexibly explore the association between NT-proBNP levels and heart failure hospitalization. This approach captures the inherent nonlinearity in biomarker–outcome relations and provides clinically interpretable cutoff values. Third, the parallel evaluation of outpatient and emergency cohorts provided a unique opportunity to compare biomarker behavior across different clinical contexts, enhancing the translational applicability of our findings.

Nonetheless, a number of limitations warrant consideration. This was a hospital-based retrospective cohort study, and the patient population may represent individuals with more advanced or complex diabetes, which could affect the generalizability of our findings. Although left ventricular ejection fraction was available for classification of HFpEF and HFrEF, detailed echocardiographic parameters were incomplete, preventing comprehensive evaluation of cardiac structure. NT-proBNP was measured only once at baseline; longitudinal changes in biomarker levels could not be examined. Finally, while our study provides valuable evidence for ethnicity- and setting-specific NT-proBNP thresholds in Asian patients with T2D, prospective multicenter validation is warranted before clinical implementation.

To conclude, in this large real-world Asian cohort with T2D, we identified setting-specific and kidney function–adjusted NT-proBNP thresholds that predict heart failure hospitalization earlier and at lower levels than current guideline cutoffs. These findings highlight substantial ethnic and cardiorenal influences on natriuretic peptide interpretation and support recalibration of NT-proBNP thresholds to improve early heart failure detection in Asian populations.

## Data Availability

The data supporting this study's findings are not publicly available due to patients privacy but are available from the corresponding author upon reasonable request and approval by the Institutional Review Boards.

## Acknowledgments

The authors are grateful to the staff of the Department of Medical Research for providing clinical data from the National Taiwan University Hospital-integrative Medical Database.

## Supplemental Material

Tables S1

Figure S1-S8

## Abbreviations

NT-proBNP: N-terminal pro-B-type natriuretic peptide
CKD: chronic kidney disease
BMI: body mass index
T2D: type 2 diabetes
eGFR: estimated glomerular filtration rate
HTN: hypertension
CHD: coronary heart disease
CVD: cerebrovascular disease
GAM: generalized additive model

## References

1. Khan MAB, Hashim MJ, King JK, Govender RD, Mustafa H, Al Kaabi J. Epidemiology of Type 2 Diabetes - Global Burden of Disease and Forecasted Trends. J Epidemiol Glob Health. 2020;10:107–111. doi: 10.2991/jegh.k.191028.001

2. van Dieren S, Beulens JW, van der Schouw YT, Grobbee DE, Neal B. The global burden of diabetes and its complications: an emerging pandemic. Eur J Cardiovasc Prev Rehabil. 2010;17 Suppl 1:S3–8. doi: 10.1097/01.hjr.0000368191.86614.5a

3. McAllister DA, Read SH, Kerssens J, Livingstone S, McGurnaghan S, Jhund P, Petrie J, Sattar N, Fischbacher C, Kristensen SL, et al. Incidence of Hospitalization for Heart Failure and Case-Fatality Among 3.25 Million People With and Without Diabetes Mellitus. Circulation. 2018;138:2774–2786. doi: 10.1161/circulationaha.118.034986

4. Dunlay SM, Givertz MM, Aguilar D, Allen LA, Chan M, Desai AS, Deswal A, Dickson VV, Kosiborod MN, Lekavich CL, et al. Type 2 Diabetes Mellitus and Heart Failure: A Scientific Statement From the American Heart Association and the Heart Failure Society of America: This statement does not represent an update of the 2017 ACC/AHA/HFSA heart failure guideline update. Circulation. 2019;140:e294–e324. doi: 10.1161/cir.0000000000000691

5. Shah AD, Langenberg C, Rapsomaniki E, Denaxas S, Pujades-Rodriguez M, Gale CP, Deanfield J, Smeeth L, Timmis A, Hemingway H. Type 2 diabetes and incidence of cardiovascular diseases: a cohort study in 1·9 million people. Lancet Diabetes Endocrinol. 2015;3:105–113. doi: 10.1016/s2213-8587(14)70219-0

6. Verma S, Sharma A, Kanumilli N, Butler J. Predictors of heart failure development in type 2 diabetes: a practical approach. Curr Opin Cardiol. 2019;34:578–583. doi: 10.1097/hco.0000000000000647

7. Mueller C, McDonald K, de Boer RA, Maisel A, Cleland JGF, Kozhuharov N, Coats AJS, Metra M, Mebazaa A, Ruschitzka F, et al. Heart Failure Association of the European Society of Cardiology practical guidance on the use of natriuretic peptide concentrations. Eur J Heart Fail. 2019;21:715–731. doi: 10.1002/ejhf.1494

8. Wong CW, Tafuro J, Azam Z, Satchithananda D, Duckett S, Barker D, Patwala A, Ahmed FZ, Mallen C, Kwok CS. Misdiagnosis of Heart Failure: A Systematic Review of the Literature. J Card Fail. 2021;27:925–933. doi: 10.1016/j.cardfail.2021.05.014

9. Januzzi JL, van Kimmenade R, Lainchbury J, Bayes-Genis A, Ordonez-Llanos J, Santalo-Bel M, Pinto YM, Richards M. NT-proBNP testing for diagnosis and short-term prognosis in acute destabilized heart failure: an international pooled analysis of 1256 patients: the International Collaborative of NT-proBNP Study. Eur Heart J. 2006;27:330–337. doi: 10.1093/eurheartj/ehi631

10. Weber M, Hamm C. Role of B-type natriuretic peptide (BNP) and NT-proBNP in clinical routine. Heart. 2006;92:843–849. doi: 10.1136/hrt.2005.071233

11. Heidenreich PA, Bozkurt B, Aguilar D, Allen LA, Byun JJ, Colvin MM, Deswal A, Drazner MH, Dunlay SM, Evers LR, et al. 2022 AHA/ACC/HFSA Guideline for the Management of Heart Failure: A Report of the American College of Cardiology/American Heart Association Joint Committee on Clinical Practice Guidelines. Circulation. 2022;145:e895–e1032. doi: 10.1161/cir.0000000000001063

12. McDonagh TA, Metra M, Adamo M, Gardner RS, Baumbach A, Böhm M, Burri H, Butler J, Čelutkienė J, Chioncel O, et al. 2021 ESC Guidelines for the diagnosis and treatment of acute and chronic heart failure. Eur Heart J. 2021;42:3599–3726. doi: 10.1093/eurheartj/ehab368

13. Marx N, Federici M, Schütt K, Müller-Wieland D, Ajjan RA, Antunes MJ, Christodorescu RM, Crawford C, Di Angelantonio E, Eliasson B, et al. 2023 ESC Guidelines for the management of cardiovascular disease in patients with diabetes. Eur Heart J. 2023;44:4043–4140. doi: 10.1093/eurheartj/ehad192

14. Su TH, Yang SS, Lee MH, Kao WY, Huang SC, Chen FF, Poon FS, Tsai LW, Chen YT, Lin C, et al. High Steatosis-Associated Fibrosis Estimator scores predict hepatocellular carcinoma in viral and non-viral hepatitis and metabolic dysfunction-associated steatotic liver disease. Clin Mol Hepatol. 2025;31:796–809. doi: 10.3350/cmh.2024.0822

15. Levey AS, Stevens LA, Schmid CH, Zhang YL, Castro AF, 3rd, Feldman HI, Kusek JW, Eggers P, Van Lente F, Greene T, et al. A new equation to estimate glomerular filtration rate. Ann Intern Med. 2009;150:604–612. doi: 10.7326/0003-4819-150-9-200905050-00006

16. Gupta DK, Claggett B, Wells Q, Cheng S, Li M, Maruthur N, Selvin E, Coresh J, Konety S, Butler KR, et al. Racial differences in circulating natriuretic peptide levels: the atherosclerosis risk in communities study. J Am Heart Assoc. 2015;4. doi: 10.1161/jaha.115.001831

17. McKie PM, Cataliotti A, Lahr BD, Martin FL, Redfield MM, Bailey KR, Rodeheffer RJ, Burnett JC, Jr. The prognostic value of N-terminal pro-B-type natriuretic peptide for death and cardiovascular events in healthy normal and stage A/B heart failure subjects. J Am Coll Cardiol. 2010;55:2140–2147. doi: 10.1016/j.jacc.2010.01.031

18. Tsutsui H, Isobe M, Ito H, Ito H, Okumura K, Ono M, Kitakaze M, Kinugawa K, Kihara Y, Goto Y, et al. JCS 2017/JHFS 2017 Guideline on Diagnosis and Treatment of Acute and Chronic Heart Failure - Digest Version. Circ J. 2019;83:2084–2184. doi: 10.1253/circj.CJ-19-0342

19. Lee KH, Kim JY, Koh SB, Lee SH, Yoon J, Han SW, Park JK, Choe KH, Yoo BS. N-Terminal Pro-B-type Natriuretic Peptide Levels in the Korean General Population. Korean Circ J. 2010;40:645–650. doi: 10.4070/kcj.2010.40.12.645

20. McDonagh TA, Metra M, Adamo M, Gardner RS, Baumbach A, Böhm M, Burri H, Butler J, Čelutkienė J, Chioncel O, et al. 2021 ESC Guidelines for the diagnosis and treatment of acute and chronic heart failure: Developed by the Task Force for the diagnosis and treatment of acute and chronic heart failure of the European Society of Cardiology (ESC). With the special contribution of the Heart Failure Association (HFA) of the ESC. Eur J Heart Fail. 2022;24:4–131. doi: 10.1002/ejhf.2333

21. Parlati ALM, Madaudo C, Nuzzi V, Manca P, Gentile P, Di Lisi D, Jordán-Ríos A, Shamsi A, Manzoni M, Sadler M, et al. Biomarkers for Congestion in Heart Failure: State-of-the-art and Future Directions. Card Fail Rev. 2025;11:e01. doi: 10.15420/cfr.2024.32

22. Mishra RK, Li Y, Ricardo AC, Yang W, Keane M, Cuevas M, Christenson R, deFilippi C, Chen J, He J, et al. Association of N-terminal pro-B-type natriuretic peptide with left ventricular structure and function in chronic kidney disease (from the Chronic Renal Insufficiency Cohort [CRIC]). Am J Cardiol. 2013;111:432–438. doi: 10.1016/j.amjcard.2012.10.019

23. DeFilippi CR, Fink JC, Nass CM, Chen H, Christenson R. N-terminal pro-B-type natriuretic peptide for predicting coronary disease and left ventricular hypertrophy in asymptomatic CKD not requiring dialysis. Am J Kidney Dis. 2005;46:35–44. doi: 10.1053/j.ajkd.2005.04.007

24. van Kimmenade RR, Januzzi JL, Jr., Baggish AL, Lainchbury JG, Bayes-Genis A, Richards AM, Pinto YM. Amino-terminal pro-brain natriuretic Peptide, renal function, and outcomes in acute heart failure: redefining the cardiorenal interaction? J Am Coll Cardiol. 2006;48:1621–1627. doi: 10.1016/j.jacc.2006.06.056

25. Chen F, Li JQ, Ou YW, Xia TL, Huang FY, Chai H, Huang BT, Li Q, Pu XB, Li GY, et al. The impact of renal function on the prognostic value of N-terminal pro-B-type natriuretic peptide in patients with coronary artery disease. Cardiol J. 2019;26:696–703. doi: 10.5603/CJ.a2018.0031

26. Ma H, Zhou J, Zhang M, Shen C, Jiang Z, Zhang T, Gao F. The Diagnostic Accuracy of N-Terminal Pro-B-Type Natriuretic Peptide and Soluble ST2 for Heart Failure in Chronic Kidney Disease Patients: A Comparative Analysis. Med Sci Monit. 2023;29:e940641. doi: 10.12659/msm.940641

27. Srisawasdi P, Vanavanan S, Charoenpanichkit C, Kroll MH. The effect of renal dysfunction on BNP, NT-proBNP, and their ratio. Am J Clin Pathol. 2010;133:14–23. doi: 10.1309/ajcp60htpgigfcnk

